# Can herd immunity be achieved without breaking ICUs?

**DOI:** 10.1101/2020.05.26.20113746

**Authors:** Edward De Brouwer, Daniele Raimondi, Yves Moreau

**Affiliations:** ESAT-STADIUS, KU Leuven, 3001 Leuven, Belgium

## Abstract

The current COVID-19 pandemic led to the rapid overload of Intensive Care Units (ICUs) in countries where the outbreaks was not quickly controlled.

The containment measures put in place to control the outbreaks had a huge social and economic impacts, and countries are looking for strategies to relax these measures while maintaining the *R*_0_ close or below 1, in an attempt to safely reach herd immunity.

In this paper we analyse the feasibility of reaching herd immunity without saturating ICUs across countries. We provide an online tool, available at www.about-the-curve.net that simulates the time required for such a scenario with a SIR model. For United States, we find that a minimum of 5 months would be required, 22 months for UK, 1 year for Italy and 9 months for Belgium.

**Disclaimer:** The presented results are preliminary and have not been peer-reviewed.

---

A key aspect of the current COVID-19 pandemic has been the rapid overload of Intensive Care Units (ICUs) in countries and regions where the epidemic was not quickly controlled [1, 2] because many patients infected with SARS-CoV-2 develop Acute Respiratory Distress Syndrome (ARDS) and need respiratory support [1].

While mortality currently appears significant in high-risk groups, it seems fairly low outside those groups [3], with a Case Fatality Rate (CFR) higher for patients > 60 yrs old and with comorbidities, such as hypertension (odds ratio 2.36 (95% CI: 1.46 to 3.83)), respiratory disease (2.46 (1.76 to 3.44)), and cardiovascular disease (3.42 (1.88 to 6.22)) [4, 3].

The relatively low mortality outside the high-risk group, the absence of a vaccine and of an antiviral treatment, and the huge socioeconomic impact of the pandemic (which will itself cause significant mortality, morbidity, psychological distress, and economic suffering) suggest that strategies aiming at achieving herd immunity, even at the cost of substantial mortality, could be evaluated. Once herd immunity has been achieved, high-risk individuals are protected. By contrast, strategies based on containment are inherently fragile as temporary failure to maintain the effective reproduction number *R* below 1 leads to the flare-up of the disease.

The outbreaks in several major cities or regions (Wuhan, Lombardy, New York, France’s Grand Est region, etc.) have shown that letting the disease run its course, even for a few weeks, leads to the rapid oversaturation of ICUs [5]. As a result, many countries, regions, and cities have had to resort to draconian lockdown measures, which have brought outbreaks under control after about 3 weeks with a plateau in hospitalizations followed by a later decrease. However, such a drop only results from the severe lockdown measures with harsh social and economic consequences. The complete lifting of those lockdown measures without appropriate alternative measures would automatically lead to a dramatic rebound of the epidemic. As such, locations currently under lockdown are partly cornered by this choice as they search for strategies to prevent a rebound when they lift lockdown.

In the classical Susceptible-Infected-Recovered model [6], herd immunity is achieved when a fraction (1 − 1/*R*_0_) of the population is immune against the disease and the height of the epidemic peak is given by (1 − 1/*R*_0_ − ln(*R*_0_)/*R*_0_) in an immunologically naive population (no individual immune at the start of the epidemic). For example, for *R*_0_ = 2.2 [7], herd immunity would be achieved if 55% of the population is immune, and 19% of the population would be infectious at the epidemic peak. Although it is still unclear which fraction of the population requires ICU care following SARS-CoV-2 infection, the figure of 2% can be used as an estimate [5, 8] for populations with a constrictive (“older”) age pyramid (current reports of 5% are for laboratory confirmed cases, which is likely to miss mild and asymptomatic cases). If correct, this would imply that 4 in 1,000 people would require ICU care at the epidemic peak, while preexisting ICU bed capacity in high-income countries [9] ranges from 4.2 per 100,000 inhabitants (Portugal) to 34.7 per 100,000 (United States). (Note that Turkey’s ICU bed capacity is 46 per 100,000.) Even if the SIR model only gives a crude estimate, it clearly shows that baseline ICU capacity and possible peak need are simply on different scales.

So, to avoid oversaturation of ICU capacity, one could imagine pushing down the effective reproduction number *R*. In fact, an *R* that does not lead to oversaturation of ICU capacity would have to be very close to 1. Under the SIR model, an ICU bed capacity of 10 per 100,000 inhabitants and 2% of infected individuals requiring ICU care, *R* would have to be around 1.1. A more practical setup would be bringing the ICU load to a fixed capacity that the local healthcare system can handle over the longterm by and then “work through the population” until herd immunity is achieved. This is essentially the position currently defended by some countries (Netherlands [10], Sweden[11]): avoid overburdening the ICU, but let the disease move forward in a controlled manner to build the immunity of the population and eventually achieve herd immunity. Note that this would require that *R* equal 1 on average.

Such an approach would however raise two important questions. First, how much time would be required to achieve herd immunity? Second, how do you control the number of (ICU) cases at a fixed level. We tackle here the first question by providing an online app (www.about-the-curve.net) that allows running different scenarios and estimating the period needed to achieve herd immunity. Concerning the period of time needed to achieve herd immunity, two important remarks need to be made. First, if this period is longer than the time needed to develop a vaccine, then the usefulness of the strategy is questionable. Second, if this period is longer than the duration of the immunity, part of the individuals who had become immune will have become susceptible again, so that herd immunity might be completely out of reach. There is considerable uncertainty about the duration of immunity following COVID-19 recovery. Immunity has been documented to be around 18 months to 2 years following SARS recovery [12]. This has been considered the default scenario. However, some coronaviruses (HCoV-OC43, HCoV-HKU1) generate little immunity [13]. There have been several reports of reinfections following COVID-19 recovery and the issue is still debated [14, 15, 16], since it is unclear whether such reports could have been the results of false negative diagnostic results [17]. Our web application allows computing the period for reaching herd immunity taking uncertainty into account.

To illustrate the model, we can consider a range of scenarios for the United States (331 million inhabitants) with a baseline ICU capacity of 34.7 beds per 100,000 inhabitants (= 113,000 beds). Assuming *R*_0_ between 2 and 3, a COVID ICU capacity between 15 and 30 beds per 100,000 inhabitants (or between 49,000 and 98,000 beds), an average ICU stay duration between 10 and 15 days, and a percentage of ICU admission following SARS-CoV-2 infection in the general population between 0.7% and 1.5%, we obtain a time to herd immunity ranging between 5 and 18 months. Assuming a fatality rate among ICU patients ranging between 33% and 50%, we obtain a number of COVID fatalities at ICUs ranging between 497,000 and 1,273,000. While the time to herd immunity could be acceptable in the best case scenario, it could be equal to the time necessary to develop a vaccine and/or the average duration of the immunity in the worst-case scenario. By contrast, taking the case of the United Kingdom with a baseline capacity of 6.6 beds per 100,000 inhabitants, assuming a long-term COVID ICU bed capacity between 5 and 10 beds per 100,000 inhabitants, and keeping all the other parameters the same as for the US scenarios, the range for the time to immunity would be between 16 and 55 months. Obviously, all parameters are open for discussion, so that the application does not return one specific prediction, but rather a range for estimates for a range of scenarios. Note that if the user wishes to consider a single value for a given parameter, both limits of the range can simply be set to be equal.

Regarding the second question raised by keeping the ICU load constant, we note that such strategies would require tightly controlling the effective reproduction number *R_e_* (the equivalent of *R*_0_ when measures are implemented to control the epidemic, which may vary over time) on average at 1. Whenever *R_e_* is above 1, the epidemic will flare up, which will quickly overload a healthcare system already at saturation. Whenever *R_e_* is below 1, the disease starts vanishing, thereby extending the time needed to achieve herd immunity. Given that it is unclear what the precise impact of any containment measure is on *R*_e_, a strategy based on lifting and reimposing measures to switch between *R_e_* slightly below 1 and *R_e_* slightly above 1 could be challenging. Moreover, constantly piloting *R_e_* on demand would require frequent changes of the NPIs imposed on the community, which might be socially infeasible.

If it turned out that the percentage of the population that has recovered from the disease asymptomatically or with minimal symptoms is significantly larger than expected, this would bring the percentage *C* of complications significantly lower and could significantly decrease the period needed to achieve herd immunity. The availability of a medical treatment that would greatly decrease the chances of complications among symptomatic patients would have a similar effect. An effective prophylactic pharmaceutical treatment [18], if only used among high-risk individuals would also have a similar effect.

## Methods

The first parameter is the *R*_0_ of COVID-19 in a population without containment measures. This allows calculating the fraction of the population 1 − 1/*R*_0_ needed to reach herd immunity. Given that *R*_0_ is uncertain, the application takes a range 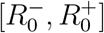 as input. This allows calculating a range of herd immunity thresholds [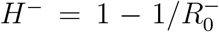; 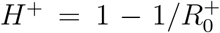] (in percent of the population) and considers these values as an error measurement: *H*^−^ = *H* −*ε_H_*; *H*^+^ = *H* +*ε_H_*. If some containment measures (hygienic measures, some level of physical distancing, such as no handshake) were to be maintained after the immunity building period, the value of *R*_0_ used could be lowered, although it is difficult to quantify the effect of such measures.

Next to the herd immunity target, we also need the ICU bed capacity in beds per 100,000 inhabitants. This is not the baseline ICU capacity in the population considered [9], but the ICU capacity fully dedicated to COVID-19 patients over the course of multiple months [5]. This figure should take into account (1) the ICU bed capacity that can be additionally deployed in a given population, (2) that the current deprioritization of all non-urgent ICU capacity is difficult to maintain in the long term, and (3) that constantly running ICUs at full capacity would make it extremely difficult to deal with other mass emergencies (such as natural or industrial disasters). This ICU bed capacity is input as an error range [*B*^−^ = *B − ε_B_*; *B*^+^ = *B* + *ε_B_*] (in beds per 100,000 inhabitants). The next variable is the average duration at ICU as this determines the average number of daily admissions because the number of ICU COVID patients needs to stay constant. Because there is a large difference in length of stay for fatalities vs. survivors, we use two ranges: [*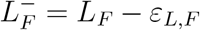*; 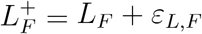] (in days for fatalities) and [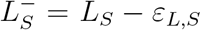; 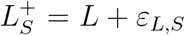] (in days for survivors). We also then require a range for the ICU fatality rate F (in percent): [*F*^−^ = *F* − *ε_F_*; *F* + = *F* + *ε_F_*]. This allows computing a range for the length of stay across all ICU patients as [*L*^−^ = *L* − *ε_L_*; *L*+ = *L* + *ε_L_*] with *L* = *F.L_F_* + (1 − *F*).*L_S_* and

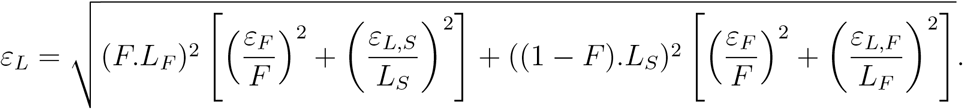

Next, we need the fraction of the population that will require ICU care following SARS-CoV-2, which is the most uncertain factor. The fraction of the general population that recovers from SARS-CoV-2 infection asymptomatically or with only mild symptoms is poorly characterized at this point because testing efforts have focused on symptomatic cases and/or people who have been in contact with infectious patients. Systematic serological surveys are needed to reduce the uncertainty for this factor. This fraction is also input as a range [*C^−^* = *C − ε_C_*; *C*^+^ = *C* + *ε_C_*] (in percent of the population).

Strategies that aim at segregating high-risk patients from the rest of the population could use a lower fraction, with the aim at achieving a level of immunity in the low-risk population sufficient to guarantee herd immunity for the total population. It is important to note that if the population-wide herd immunity threshold is given by 1 − 1/*R*_0_, and a proportion *P* of the population remains segregated, then the immunization rate that needs to be achieve in the exposed population needs to be increased to (*R*_0_ *−* 1)/(*R*_0_.(1 − *P*)).

The average number of days till herd immunity can then be computed as

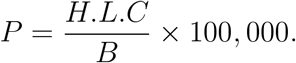

The error range can be calculated using error analysis:

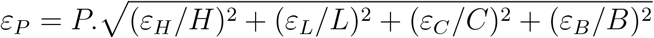

and a range can be returned as [*P*^−^ = *P* − *ε_P_*; *P* + = *P* + *ε_P_*].

We can also consider further the number of fatalities among ICU COVID patients in such a strategy. Ultimately, the percentage of the population that will have been infected will be in the range [*H*^−^; *H*+], the fraction of the population needed to achieve herd immunity. The range for the percentage of fatalities in the population is then given by *D* = *H.F*, 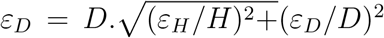, and the range [*D^−^* = *D − ε_D_*; *D*^+^ = *D* + *ε_D_*].

## Data Availability

All data was collected from :https://coronavirus.jhu.edu/map.html

## Funding

YM is funded by Research Council KU Leuven: C14/18/092 SymBioSys3; CELSA-HIDUCTION CELSA/17/032 Flemish Government:IWT: Exaptation, PhD grants FWO 06260 (Iterative and multi-level methods for Bayesian multirelational factorization with features) This research received funding from the Flemish Government under the “Onderzoeksprogramma Artificiele Intelligentie (AI) Vlaanderen” programme. EU: “MELLODDY” This project has received funding from the Innovative Medicines Initiative 2 Joint Undertaking under grant agreement No 831472. This Joint Undertaking receives support from the European Union’s Horizon 2020 research and innovation programme and EFPIA. DR is funded by a FWO postdoctoral fellowship and EDB is funded by a FWO-SB grant.

## Competing interests

The authors declare no competing interests.

## Notes

### Competing Interest Statement

The authors have declared no competing interest.

### Author Declarations

No IRB or ethics committee required.

